# Erythrocytes Reveal Complement Activation in Patients with COVID-19

**DOI:** 10.1101/2020.05.20.20104398

**Authors:** LK Metthew Lam, Sophia J. Murphy, Leticia Kuri-Cervantes, Ariel R. Weisman, Caroline A. G. Ittner, John P. Reilly, M. Betina Pampena, Michael R Betts, E. John Wherry, Wen-Chao Song, John D. Lambris, Douglas B. Cines, Nuala J. Meyer, Nilam S. Mangalmurti

## Abstract

COVID-19, the disease caused by the SARS-CoV-2 virus, can progress to multi-organ failure characterized by respiratory insufficiency, arrhythmias, thromboembolic complications and shock ^1-5^. The mortality of patients hospitalized with COVID-19 is unacceptably high and new strategies are urgently needed to rapidly identify and treat patients at risk for organ failure. Clinical epidemiologic studies demonstrate that vulnerability to organ failure is greatest after viral clearance from the upper airway ^6-8^, which suggests that dysregulation of the host immune response is a critical mediator of clinical deterioration and death. Autopsy and pre-clinical evidence implicate aberrant complement activation in endothelial injury and organ failure ^9,10^. A potential therapeutic strategy warranting investigation is to inhibit complement, with case reports of successful treatment of COVID-19 with inhibitors of complement ^10-13^. However, this approach requires careful balance between the host protective and potential injurious effects of complement activation, and biomarkers to identify the optimal timing and candidates for therapy are lacking. Here we report the presence of complement activation products on circulating erythrocytes from hospitalized COVID-19 patients using flow cytometry. These findings suggest that novel erythrocyte-based diagnostics provide a method to identify patients with dysregulated complement activation.

## Introduction

An ancient arm of innate immunity, the complement system provides a front-line of defense against invading micro-organisms. This multi-tiered and highly coordinated system is vital for the innate immune response to pathogens, removal of dead cells and maintenance of homeostasis. Initially described as a complement to antibody mediated immunity, it was later discovered that the complement system is evolutionarily older than antibody-mediated immunity and its activation can be initiated by lectins and by a C3 tick-over mechanism. Thus, the three pathways of complement activation can be engaged by distinct initiators including antigen-antibody complexes (classical pathway), lectins (lectin pathway) and spontaneous C3 hydrolysis (alternative pathway). Although the complement system promotes clearance of pathogens through opsonization, inflammation and cytolysis, dysregulated complement activation can lead to cellular injury, microvascular thrombosis and organ failure ^14^ The latter point is highlighted by several well-characterized human diseases including paroxysmal nocturnal hemoglobinuria (PNH) and atypical hemolytic uremic syndrome (aHUS) that result from complement dysregulation ^15,16^. Animal models with complement regulator deficiencies or mutations have also demonstrated development of tissue injury, coagulopathy and thrombotic microangiopathy caused by insufficient complement regulation ^17^

Complement-containing immune complexes bind to cells through a number of specific receptors, including complement receptor 1 (CR1), which recognizes complement activation products C3b and iC3b ^18^. Human CR1 is abundantly expressed on the surface of the nearly thirty trillion erythrocytes (red blood cells, RBCs) in circulation. Immune adherence, binding of antigen-antibody-complement complex to RBCs, was first described in the 1950s and is one example of how erythrocytes modulate innate responses ^19-21^. We hypothesized that complement deposition on circulating RBCs would provide a sensitive measure to detect complement activation that may be occurring in hospitalized patients with COVID-19.

## Results and Discussion

To detect complement activation in patients with COVID-19, we measured erythrocyte-bound C3b, iC3b, C3dg and C4d using flow cytometry (see online methods). RBCs were obtained from healthy donors (HD) or patients with COVID-19 (Table 1, online methods) on day 0 and day 7 of study enrollment. The percentage of RBCs with bound C3b/iC3b/C3dg was markedly elevated in hospitalized COVID-19 patients admitted to the ICU when compared with HD and increased even further by day 7 (Figure 1 a&b). C4d was also increased on RBCs from COVID patients when compared with healthy donors (Supplemental Figure 1). Immunofluorescence staining demonstrated that COVID-19 erythrocytes not only bound C3 fragments, but also bound viral spike protein, suggesting activation of the classic pathway of complement and immune complex deposition on the RBCs (Fig 2). Together, these data suggest that complement activation products and viral antigen are present on RBCs in patients with COVID-19.

**Supplemental Figure 1.**
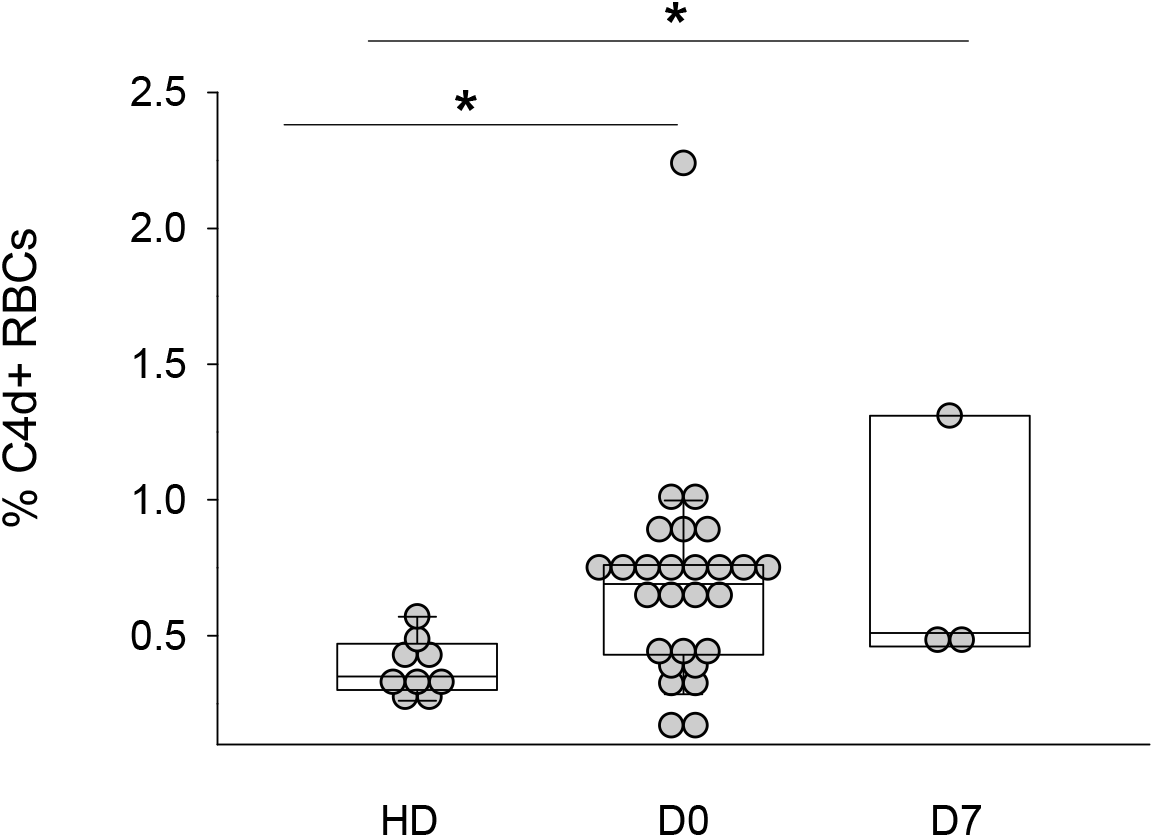
**C4d is bound to COVID patient RBCs**. Flow cytometry was performed on RBCs obtained from healthy donors (HD) or critically ill COVID-19 patients on the day of ICU admission (D0) and 7 days after ICU admission (D7), data is expressed as percent RBCs that have bound C4d. *p=0.016 by one-way ANOVA, n=9 HD, n=27 D0, n=3, D7.

We next examined the association of RBC-bound complement, C-reactive protein (CRP), d-dimer and clinical outcomes (patient details are in Table 1). We detected a moderate inverse correlation (rho −0.55) between the percentage of C3 bearing RBCs at the early timepoint and the clinically measured high sensitivity CRP (p=0.05), measured in 13/35 subjects, but not with the duration of symptoms before blood draw, d-dimer level, absolute lymphocyte count, ferritin level, or maximum oxygenation required. The median C3 percentage was not different between subjects who developed ARDS or not, though observations are limited and inconclusive given the small cohort of patients studied.

The presence of C4 and C3 on patient RBCs suggests activation of the classical pathway of complement. Direct binding of the N protein of SARS-CoV-1 or SARS-CoV-2 to MASP-2 (Mannan-binding lectin serine protease 2) may also lead to activation of the lectin pathway as well ^10^. Polymorphisms in the MBL protein, which initiates the lectin pathway, and mutations or dysfunction of complement regulatory proteins are implicated in a diverse group of disorders characterized by intravascular thrombosis ^15-17,22-24^. Furthermore, risk factors for COVID-associated lung injury such as diabetes have been associated with high circulating levels of MBL and dysfunctional complement regulatory proteins on the endothelium and erythrocytes ^25-29^.

**Figure 1.**
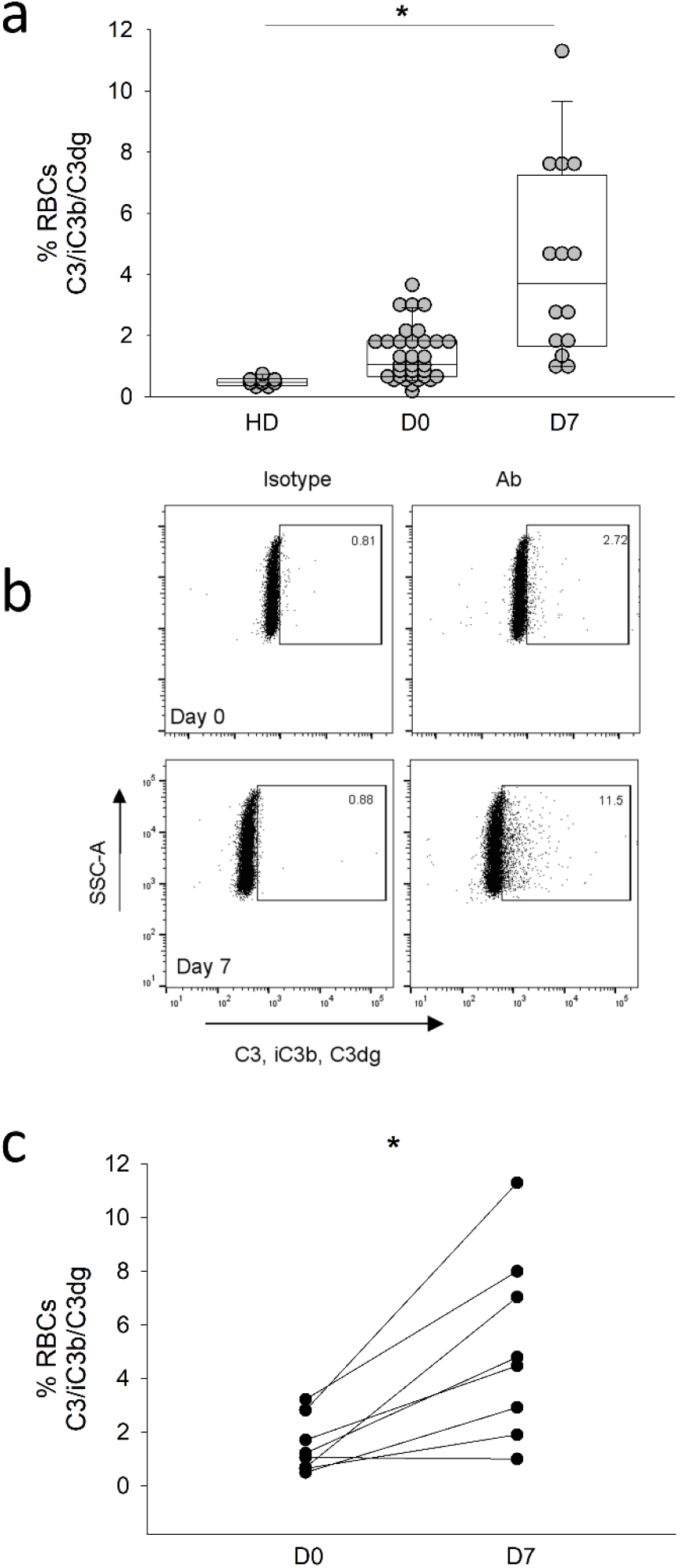
**C3 fragments are bound to COVID patient RBCs**. a. Flow cytometry was performed on RBCs obtained from healthy donors (HD) or critically ill COVID-19 patients on the day of ICU admission (D0) and 7 days after ICU admission (D7). Data are expressed as percent RBCs that have bound C3. *p<0.001 one-way ANOVA, pairwise comparisons by Dunn’s test: HD v D0 *p=0.007, HD v D7 *p<0.001, D7 v D0 *p=0.003. n=9, 33 and 14 for HD, D0 and D7. b. Representative dot plot from a critically ill patient D0 and D7. c. C3 fragments are increased over time in critically ill COVID-19 patients, *p=0.007 paired t-test, *p=0.01 by Rank Sum test, n=8.

**Figure 2.**
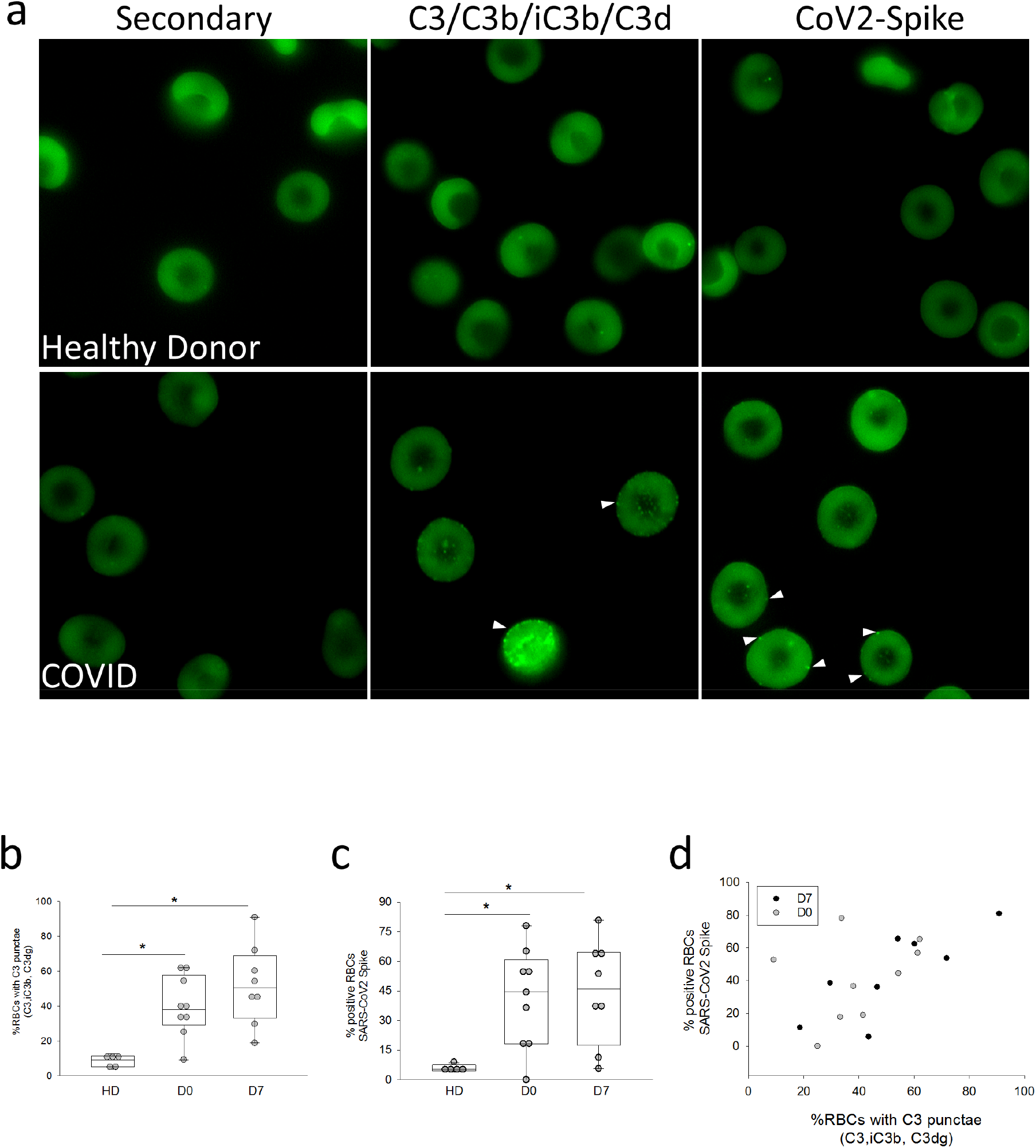
**Spike protein and complement fragments are bound to RBCs**. a. Immunofluorescence of RBCs from HD and critically ill COVID-19 patients. White arrowheads denote Spike protein punctae. b. Quantification of C3 punctae, *p= 0.002 by ANOVA. p=0.01 HD v D0, p=0.001 HD v D7, p=NS D0 v D7 by multiple comparisons (Holm-Sidak). C. Quantification of SARS-CoV2 Spike protein, *p=0.02 by ANOVA. p=0.049 HD v D0, p=0.026 HD v D7, p=NS D0 v D7 by multiple comparisons (Holm-Sidak). n=5-10 patient samples, 3-5 fields analyzed/sample. Correlation between RBC-bound Spike protein and RBC-bound C3 fragments, correlation coefficient =0.59, *p=0.01. D7 correlation coefficient =0.078, *p=0.014. D0=NS.

**Table 1.** Baseline Characteristics of the COVID-19 patients and healthy donors.

|  | Healthy Donors<br>(n=9) | COVID-19 Inpatients<br>(n=35) |
| --- | --- | --- |
| Age | 36 ± 8 | 61 ± 15 |
| Male | 4 (40%) | 19 (54%) |
| Race |  |  |
| African American | 0 (0%) | 19 (54%) |
| White | 8 (89%) | 11(31%) |
| Asian | 1 (11%) | 5 (14%) |
| APACHE III score | NA | 65 (52 - 80) |
| D-dimer at blood draw | NA | 1.25 (0.72 – 4.36) |
| C reactive protein, high sensitivity | NA | 134 (97 - 160) |
| Absolute lymphocyte count at blood draw | NA | 0.8 (0.6 – 1.5) |
| Severity of pneumonia* | NA |  |
| Moderate |  | 9 (26%) |
| Severe |  | 26 (74%) |
| Maximum FiO2 applied (%) | NA | 60 (30 – 100) |
| ARDS | NA | 16 (46%) |
| Septic shock | NA | 15 (44%) |
| Clotting complication | NA | 11 (32%) |
| Mortality | NA | 5 (15%) |

\*Moderate: oxygen ≤ 6L NC; Severe: oxygen by high flow nasal cannula, non-invasive ventilation, or invasive mechanical ventilation

Although the small sample size did not permit us to draw associations between RBC-bound complement and clinical outcomes, several clinically relevant implications arise from our findings. First, our findings suggest support a potential role for complement in the pathophysiology of COVID-19. Second, because plasma-based assays of individual complement components or total complement activity may fail to reflect the levels of complement activation on cell surfaces that is responsible for tissue injury ^30^, flow cytometry of RBCs may serve as a readily accessible and sensitive measure of complement activation. Third, an increase in RBC-bound complement over time might serve as a biomarker for severe COVID-19 disease and help to depict disease trajectory and identify potential patients for clinical trials of anti-complement therapy.

Interfering with complement activation requires the need to balance the protective and potentially injurious effects of these proteins on the host. The evolutionarily ancient origins of this system coupled with its importance in host defense against highly virulent bat-borne viruses including Nipah Virus, MERS-CoV and SARS-CoV-1 and 2 suggest that complement activation represents an atavistic response of the host ^10,31-33^. Moreover, immune adherence, the binding of complement-containing immune complex to erythrocytes, may provide a mechanism to immobilize and transport pathogens to immune cells ^21^. On the other hand, deposition of immune complexes and complement on RBCs may alter their rheology and thereby promote intravascular stasis and thrombosis that contribute to the pathogenesis of virus associated lung injury ^10,32,34-38^. Additional studies will be needed to determine if and when binding of complement and viral antigen to erythrocytes contributes directly to organ injury during COVID-19 infection.

In sum, our findings suggest that RBC-immune adherence occurs in COVID-19 and flow cytometric analysis of RBCs provides a clinically feasible means to help identify patients who are susceptible to complement-mediated host injury and might therefore benefit from intervention with complement inhibitors. The role of RBC immune adherence in delivery of viral antigen to immune cells has yet to be elucidated. However, exploiting this innate immune mechanism to detect pathologic complement activation may hold one key to preventing organ injury in infectious and autoimmune systemic disorders characterized by complement dysregulation.

## Methods

### SARS-CoV2/COVID-19 cohort and blood processing

Whole blood was obtained in EDTA tubes (BD Bioscience) from inpatient subjects with SARS-CoV2 positive results who were enrolled in the Molecular Epidemiology of Severe Sepsis in the ICU-COVID (MESSI-COVID) study at the University of Pennsylvania. Studies involving human subjects were approved by the University of Pennsylvania Institutional Review Board. Subjects were screened and approached for informed consent during the first 3 days of hospitalization. Informed consent, in accordance with protocols approved by the regional ethical research boards and the Declaration of Helsinki, was given by subjects or their proxies. Samples were processed within three hours of collection. Whole blood was centrifuged for 15 minutes at 3,000 rpm and plasma was removed.

#### Red blood cell isolation

Whole blood was obtained from healthy donors in EDTA tubes (BD Bioscience). Blood from healthy donors and COVID-19 patients was centrifuged for 5 minutes at 800 g and remaining plasma and buffy coat were removed. Red blood cells (RBCs) from pellet were then diluted in PBS for flow cytometry (1 uL RBCs / 1 mL PBS), confocal microscopy (1 uL RBCs / 1 mL PBS), and quantitative PCR (5 uL RBCs in 100 uL PBS). All work was done in the biosafety cabinet under enhanced-BSL2 conditions.

#### Flow cytometry

Diluted RBCs (100uL) were incubated with anti-complement C3/C3b/iC3b/C3d (BioLegend, 1 ug/mL), anti-C4d (Immunoquest, 1 ug/mL), anti-IgM (BioLegend, 5 uL), or IgG2a, K isotype (BioLegend, 1ug/mL) antibody for 45 minutes followed by two washes in PBS. Cells were then resuspended in 100 uL of PE Goat anti-mouse IgG secondary antibody (BioLegend, 2 ug/mL) for 45 minutes prior to two washes in PBS and fixation in EM grade glutaraldehyde (Polysciences, 2.5%) for 15 minutes. After subsequent washes, FACS acquisition was performed using the LSR Fortessa (BD Biosciences) and analysis was done using FlowJo Software. Data are shown as percent positive RBCs.

#### Microscopy

Diluted RBCs were fixed in EM grade glutaraldehyde (Polysciences, 0.05%) for 15 minutes prior to two washes in FACS buffer (2% FBS in PBS). Cells were then resuspended in FACS buffer (1 uL RBCs / 1 mL FACS buffer), and 100 uL of diluted RBCs were incubated with anti-complement C3/C3b/iC3b/C3d (Biolegend, 1ug/ml), anti-C4d (Immunoquest, 1 ug/mL), or anti-SARS-CoV spike (GeneTex, 1:200) antibody overnight at 4°C. Cells were subsequently washed twice in FACS buffer prior to incubation with AlexaFluor 488 Goat anti-mouse IgG secondary antibody (Thermofisher, 2 ug/mL) for 45 minutes. After subsequent washes, cells were fixed in 2.5% glutaraldehyde, washed, and mounted on slides with fluoromount G. Slides were imaged using Nikon eclipse microscope, and images were analyzed using ImageJ software. 3-5 fields/sample were counted for quantification. Data are represented as positive punctae/total RBCs.

#### Statistics

Clinical correlation were analyzed by Stata (StataCorp LLC), and all other statistical analysis were performed with SigmaPlot (SyStat Software Inc.)

### Author Contributions

Experiments were conceived and designed by NSM. Experiments were performed by SSM and ML. AW, CI, LC and MB enrolled patients and procured patient samples. MB and EJW consulted on study design. JPR and NJM enrolled and phenotyped COVID-19 patients. WCS, DBC and JDL provided vital reagents and analyzed data. Manuscript was written by NSM and DBC.

### Data and materials availability

All data is available in the main text or the supplementary materials.

## Data Availability

All data is available in the main text or the supplementary materials.

## Acknowledgements

We thank the nurses and staff at the Hospital of the University of Pennsylvania for procuring patient samples. We would also like to thank Peggy Zhang for her excellent technical assistance.

